# Bayesian Spatio-Temporal Modeling and Hotspot Mapping of Malaria Risk in Ghana

**DOI:** 10.64898/2026.05.19.26353586

**Authors:** Pascal Antwi, George Muhua, Eric Nyarko

## Abstract

**Purpose:** This study developed a Bayesian hierarchical spatio-temporal modeling framework to analyze factors and trends in malaria risk across Ghana’s 16 administrative regions from 2020 to 2024. The aim was to identify statistically significant areas with elevated or persistent malaria risk, to inform targeted intervention planning and support the National Malaria Elimination Program.

**Methods:** This study utilized malaria incidence data from the Ghana Health Service’s District Health Information Management System-II covering the years 2020 to 2024. Meteorological data were sourced from the Visual Crossing Weather Data, and regional population estimates were obtained from the Ghana Statistical Service. To analyze the data, a Bayesian hierarchical spatiotemporal model with a Negative Binomial (NB) likelihood was implemented using Integrated Nested Laplace Approximation to account for overdispersion. The model included Conditional Autoregressive priors for structured spatial effects, first-order random walk priors for temporal dependence, and spatio-temporal interaction terms. Additionally, Local Indicators of Spatial Association (LISA) analysis with 999 conditional permutations was conducted to identify statistically significant spatial clusters, including high-high hotspots and low-low cold spots.

**Results:** The NB model significantly outperformed the Poisson model, leading to a reduction in the dispersion statistic from 9,227.55 to 1.11. Humidity with a 1-month lag showed the strongest positive association with malaria risk, while the ultraviolet index had the greatest protective effect. Predictive relative risk maps identified persistent high-risk clusters in the northern and northwestern regions, specifically Upper West, Upper East, Bono, Ahafo, and Western North. LISA analysis indicated that Bono-Ahafo has been a stable high-high cluster from 2020 to 2023, while Ashanti has remained a consistent low-high anomaly. Additionally, Greater Accra and Central regions formed a significant low-low cluster in 2024.

**Conclusion:** The Bayesian hierarchical spatio-temporal framework effectively characterized the complex transmission dynamics of malaria in Ghana. It revealed significant spatial dependence, temporal correlation, and interactions between these factors. By identifying persistent high-risk clusters and statistically significant spatial associations, this framework provides essential evidence to guide resource allocation. These findings support Ghana’s National Malaria Elimination Program Strategic Plan (2024– 2028) by enabling targeted interventions in hotspots and optimizing the use of limited resources to sustain progress in low-transmission areas.

## 1 Introduction

Malaria is one of the leading causes of mortality and morbidity in developing countries and is increasingly recognized as a significant public health issue worldwide [1]. This problem is particularly severe in Southeast Asia, the Western Pacific, Latin America, and sub-Saharan Africa (SSA). In 2023, an estimated 263 million cases and 597,000 deaths due to malaria were reported, with SSA bearing the highest burden [2]. Despite a decline in national malaria prevalence—from 14.1% in 2019 to 8.6% in 2022—Ghana continues to rank among the eleven countries with the highest malaria burden in Africa, accounting for a substantial proportion of global cases and deaths [3, 4].

Ghana’s National Malaria Elimination Program Strategic Plan (2024-2028) aims to reduce malaria incidence by 50% and malaria deaths by 90% by 2028 [3]. To achieve these goals, targeted interventions informed by robust spatial risk mapping are essential, as malaria transmission dynamics exhibit significant spatial heterogeneity and temporal variability that traditional regression approaches often fail to adequately capture [5, 6]. While generalized linear models can quantify associations between predictors and disease incidence, they do not account for the inherent spatial dependence among neighboring regions or the structured temporal evolution of disease risk. These methodological limitations can lead to underestimated standard errors, inflated Type I error rates, and potentially biased conclusions about the true drivers of malaria transmission [7].

Spatial dependence arises when observations from geographically proximate areas exhibit similar values due to shared environmental characteristics, unmeasured confounders, or diffusion processes [8, 9]. In infectious disease epidemiology, this dependence is particularly pronounced in vector-borne diseases such as malaria, where factors such as mosquito populations, ecological conditions, and human movement patterns lead to natural clustering of transmission intensity [10]. The presence of spatial autocorrelation violates the independence assumption inherent in standard generalized linear models. Therefore, it is necessary to use analytical frameworks that explicitly account for spatial structure in modeling. Similarly, temporal dependence affects malaria incidence data, where case counts in one month are correlated with those in previous months due to persistent transmission cycles, delayed climatic effects, and seasonal patterns. Overlooking these temporal structures can obscure significant dynamics and jeopardize the validity of model-based inferences [11]. While lagged predictors can capture delayed effects of covariates, they do not model the underlying temporal correlation structure of the random effects, potentially leaving some temporal variation unexplained.

Spatio-temporal modeling frameworks have emerged as valuable tools for tackling these analytical challenges in disease mapping. These models decompose variation into structured spatial components, temporal trends, and their interactions, enabling more accurate estimation of disease risk while effectively quantifying uncertainty [12, 13]. The Bayesian hierarchical approach, in particular, offers a coherent probabilistic framework for integrating multiple sources of variation. This approach leverages strength across spatial units and time points, enabling stable estimates even in areas with sparse data or fluctuating case counts [14]. Within the Bayesian paradigm, Conditional Autoregressive (CAR) priors provide a natural mechanism for modeling spatial dependence by assuming that the risk in each region depends conditionally on the risks in neighboring regions [14]. Temporal dependence can be addressed using random-walk priors, which enforce smoothness over time while allowing gradual changes in risk levels [6]. The combination of these components, through a spatio-temporal interaction term, enables temporal trends to vary across space. This feature is particularly relevant for malaria, as different ecological zones experience distinct seasonal patterns and intervention effects [15].

Previous spatial analyses of malaria show significant methodological limitations. Many studies have employed Bayesian spatial and spatio-temporal models, but they assumed a Poisson distribution without formally testing for equidispersion, which may result in biased estimates or inferences [16, 17, 18, 19, 20, 21]. This study aims to address these gaps by developing a Bayesian hierarchical spatio-temporal model that utilizes a negative binomial (NB) likelihood to accommodate overdispersion, providing more reliable risk estimates for malaria control. It also explicitly models spatial dependence using CAR priors, temporal correlation through first-order random walk (RW1) priors, and their interaction. The study employs Integrated Nested Laplace Approximation (INLA) for computationally efficient Bayesian inference. Additionally, predictive relative risk maps are generated to visualize spatial and temporal patterns of malaria transmission across Ghana’s sixteen administrative regions. Finally, the study uses Local Indicator of Spatial Association (LISA) to identify statistically significant spatial clusters, including high-high hotspots, low-low cold spots, and spatial anomalies, providing detailed insights for targeted intervention planning and resource allocation.

## 2 Methods

### 2.1 Study Area and Data Source

Ghana, located in West Africa, is divided into sixteen administrative regions that span seven ecological zones, including the Sudan, Guinea, and Coastal Savannahs, as well as the Forest-Savannah Transition Zone, Semi-Deciduous Forest, and both Moist and Wet Evergreen zones. The country experiences a tropical climate characterized by distinct dry and rainy seasons, which create varied environmental conditions that influence the intensity of malaria transmission across different regions [22]. This study used malaria incidence records obtained from the Ghana Health Service’s District Health Information Management System-II (DHIMS-II). These records provide monthly regional-level totals of confirmed malaria cases from 2020 to 2024. The meteorological dataset was sourced from the Visual Crossing Weather Data [23] and includes daily records from 2020 to 2024, which were then aggregated to monthly totals. The variables in this dataset include rainfall, temperature, relative humidity, apparent temperature, dew point, sea level pressure, solar radiation, solar energy, and the ultraviolet (UV) index. Population estimates for the period from 2021 to 2024 were obtained from the Ghana Statistical Service (GSS) official population projection tables, based on the 2021 Population and Housing Census. These projections provide annual regional population estimates through 2050. Since population data for 2020 were unavailable in the official projections, we estimated regional populations for that year using linear interpolation from the projected values for 2021 and 2022, ensuring temporal consistency with the study period. To facilitate spatiotemporal modeling and spatial visualization, the administrative boundary shapefiles for Ghana’s sixteen regions were sourced from the Global Administrative Areas (GADM) spatial database. These files were obtained from the official GADM repository, and their use complies with the licensing conditions outlined on the GADM website. Ghana has two main seasons: a major and minor rainy season, separated by dry periods. This season structure is explicitly incorporated into the model.

### 2.2 Statistical Analysis

The dataset was imported into RStudio for cleaning and statistical analysis. The climatic data were aggregated to a monthly resolution by summing daily rainfall and averaging the remaining variables. Lag structures for all climatic variables were generated, with lag periods ranging from 0 to 3 months, and these lagged variables were included as covariates in the model. Since population data for 2020 were not available in the GSS official projections, the 2020 regional populations were estimated by linear interpolation from the projected values for 2021 and 2022. The interpolation was calculated as *P*_2020_ = *P*_2021_ *+* (*P*_2021_ -*P*_2022_), assuming a constant annual rate of change between consecutive years to maintain temporal consistency across the study period.

We assessed spatial dependence using global Moran’s I with Monte Carlo permutation tests (100,000 simulations), and spatial heterogeneity using Chi-square tests. A Bayesian hierarchical spatio-temporal model with a Negative Binomial likelihood was implemented to account for overdispersion. The linear predictor included lagged climatic covariates, structured spatial effects using intrinsic Conditional Autoregressive (ICAR) priors, unstructured heterogeneity, temporal effects modeled via RW1 priors, and spatio-temporal interaction terms. All parameters were assigned weakly informative priors. Estimation was performed using INLA for computational efficiency. The fundamental principle underlying INLA is that spatio-temporal models have a latent Gaussian structure that is amenable to approximation via nested Laplace methods. INLA is a fast and deterministic approach as compared to the Markov Chain Monte Carlo used in disease mapping [24]. INLA approximates posterior distributions and supports the use of a Penalized Complexity prior, which enables control of model complexity. Model comparison was performed using the Deviance Information Criterion (DIC) and the Watanabe-Akaike Information Criterion (WAIC). Posterior relative risk estimates were mapped annually, and Local Indicators of Spatial Association (LISA) analysis identified statistically significant hotspots, cold spots, and spatial outliers using local Moran’s I statistics with 999 conditional permutations. All analyses were conducted using R version 4.5.2 with the *INLA, spdep*, and *ggplot2* packages.

### 2.3 Model Specification

In this study, we implemented a Bayesian hierarchical spatio-temporal modeling framework to jointly analyze spatial and temporal dependencies in malaria incidence across Ghana’s sixteen administrative regions. This analytical approach allows us to model regional differences, temporal trends, and their interactions while incorporating climatic factors as fixed covariates. Let Ψ_*it*_ represent the observed malaria case count for region *i* (where *i =* 1, *…*,16) at time *t* (where *t= 1, …*, *T*). These counts are assumed to follow a Negative Binomial distribution, expressed as Ψ_*it*_ ∼ NB(*E*_*it*_*µ*_*it*_). Here, *E*_*it*_ denotes the expected number of cases based on national average risk, derived through indirect standardization, andµ_*it*_ represents the relative risk, which is the primary parameter of interest for inference. The linear predictor for the log-relative risk can be specified as

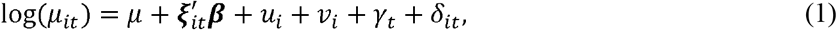

whereµ represents the overall intercept, assigned a diffuse normal prior 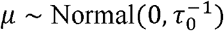. The term 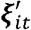 denotes the vector of lagged climatic covariates with associated coefficient vector *β*. The random effects structure comprises several components designed to capture distinct sources of unobserved variation. The term *u*_*i*_ represents spatially structured heterogeneity, modeled using CAR prior specified as

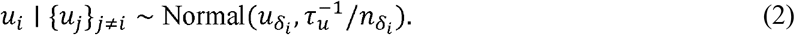

Here, 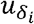 is the mean of the neighboring regions and 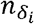 the number of neighbors for region *i*. The component *v*_*i*_ captures unstructured heterogeneity, specified as independent and identically distributed normal variates with prior 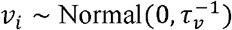. Temporal dynamics are accommodated through *y*_*t*_, which denotes temporally structured random effects modeled via RW1 expressed as

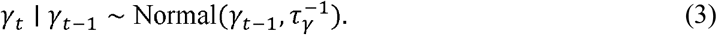

Finally, *δ*_*it*_ represents the spatio-temporal interaction term capturing residual variation not explained by the main spatial and temporal effects, with prior 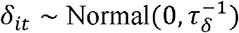. The precision parameters *τ*_*u*_, *τv, τ*_*γ*_ and *τ*_*δ*_ governing these random effect distributions are assigned Gamma hyperpriors, *τ*^*^ ∼ Gamma*(a, b)*, following [25], to reflect uncertainty in the variance components. Collectively, these priors define the latent Gaussian field underpinning the model. This hierarchical formulation flexibly accommodates spatial dependence through structured random effects, temporal evolution via autoregressive components, and local variations arising from spatio-temporal interactions in malaria transmission dynamics. The precision parameters for the random effects were assigned weakly informative Gamma priors to reflect uncertainty while allowing the data to dominate the inference. Specifically, the negative binomial dispersion parameter was assigned *τ*_*θ*_ ∼ Gamma(1,0.00005), the structured spatial effect *µ*_*i*_ had *τ*_*µ*_∼ Gamma(1,0.00005), the temporal effect *γ*_*t*_ had *τ*_*µ*_ ∼ Gamma(1,0.01), and the spatio-temporal interaction *ϕ*_*it*_ had *τ* _*ϕ*_ ∼ Gamma(1,0.00005). All regression coefficients and random effects (*β*_*0*_, ***β**,µ*_*i*_, *v*_*i*_, *γ*_*t*_, *ϕ*_*it*_) were assigned weakly informative or non-informative priors, ensuring that posterior distributions are primarily informed by the observed data.

### 2.4 Risk Mapping

The expected cases are computed as

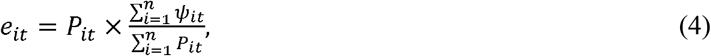

where *P*_*it*_ is the population at risk in region *i* at time *t*. The relative risk (*RR*_*it*_) in region *i* at time *t* is then computed as

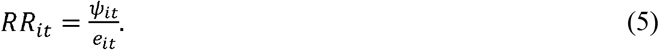

Regions and periods with *RR* > _*it*_ *1* are classified as high-risk. Posterior estimates of *RR*_*it*_ were visualized as risk maps to identify areas of elevated or reduced disease risk.

To detect hotspot malaria cases in Ghana, the study employed the LISA cluster map. A spatial approach developed by [26], the LISA map helps detect statistically significant clusters and decompose global spatial autocorrelation into specific components within a particular area. LISA enables the study to detect High-High, Low-Low, High-Low, and Low-High based on the direction and magnitude of association. The threshold to detect a hotspot area is the permutation-based pseudo *p* – *value* local Moran’s statistic given by

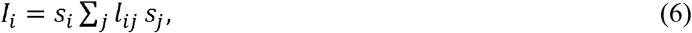

Where *s*_*i*_ and *s*_*j*_ are deviations from the mean, and *l*_*ij*_ are elements of a spatial weights matrix *L* defining neighbor relationships. This method will allow us to identify significant local patterns in the spatial distribution of malaria prevalence in Ghana. LISA maps will complement INLA-derived risk maps to visualize significant spatial patterns of malaria prevalence.

### 2.5 Metrics for Sensitivity Analysis

Sensitivity analysis assessed the robustness of spatial model estimates to alternative prior specifications. Different priors for the spatial precision parameters (*τ*_*u*_ and *τ*_*V*_) and the regression coefficients were tested to assess stability of the posterior distributions. Model comparison metrics such as DIC and WAIC will guide the evaluation of model sensitivity. The DIC can be expressed as

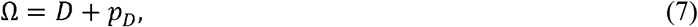

where *D* = 𝔼_*δ/ ψ*_ [*D*(*δ*)] and 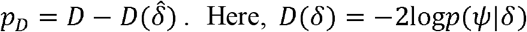 is the deviance, *D* is the posterior mean deviance, *p*_*D*_ is the effective number of parameters, and 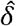 is the posterior mean of *ψ*.Lower DIC values indicate better model fit [27].

The WAIC can be expressed as

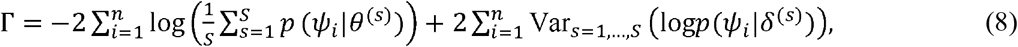

where *ψ*_*i*_ are observed data points, *δ*^(*s*)^ are posterior samples, and *s* is the total number of posterior draws. Lower WAIC values indicate better predictive accuracy [28]. Three alternative scenarios were considered: first, a stronger spatial prior, where the median precision of the structured spatial effect was reduced from

0.5 to 0.2; second, a second-order random walk (RW2) for the temporal trend, which allows for smoother temporal evolution; and third, the removal of the spatio-temporal interaction term to assess its contribution to model fit. This approach ensures reliable spatio-temporal inferences, minimizes dependence on specific prior assumptions, and improves overall model credibility.

### 2.6 Ethical Consideration

As part of the Integrated Disease Surveillance and Response, malaria cases are regularly reported in DHIMS-II. Therefore, no formal ethical approval was required for this study, as we used aggregated, de-identified secondary data for the analysis. The Ghana Health Service provided us with the dataset and granted permission to use it at no cost.

## 3 Results

### 3.1 Spatio-Temporal Model Test

In this study, we formally tested for overdispersion using the Pearson statistic. The NB model significantly outperformed the Poisson model, reducing the dispersion statistic from 9,227.55 to 1.11. We also conducted diagnostic tests for spatial dependence and heterogeneity before proceeding with spatio-temporal modeling. To assess global spatial autocorrelation, we employed Monte Carlo permutation tests using Moran’s I statistic. Additionally, we used Chi-square tests to assess regional heterogeneity in malaria incidence. Table 1 summarizes the results from the Monte Carlo permutation test for global spatial autocorrelation in the malaria incidence data. The observed Moran’s I value was 0.14516, indicating a positive clustering pattern, where regions with similar malaria incidence tend to be geographically close to one another. Based on 100,000 simulations, the observed rank of this statistic was high (91,982), yielding a p-value of 0.04018. These results confirm statistically significant positive spatial autocorrelation in malaria incidence across Ghana’s 16 regions, supporting the need to incorporate spatial effects into our analytical framework.

**Table 1.** Monte Carlo Moran’s I Test for Spatial Dependence.

| Statistic | Value |
| --- | --- |
| Observed Moran’s I | 0.14516 |
| Number of simulations | 100,000 |
| Observed rank | 91,982 |
| p-value | 0.04018 |

Table 2 shows the findings from the Chi-square test for spatial heterogeneity in malaria incidence across the 16 regions of Ghana. The test statistic was 160,839 with 15 degrees of freedom, which is exceedingly large, yielding a p-value of less than 2.2 x 10^*-16*^. This result confirms significant spatial variation in the observed malaria burden, reinforcing the necessity for models that can accommodate region-specific effects, such as structured spatial random effects.

**Table 2.**
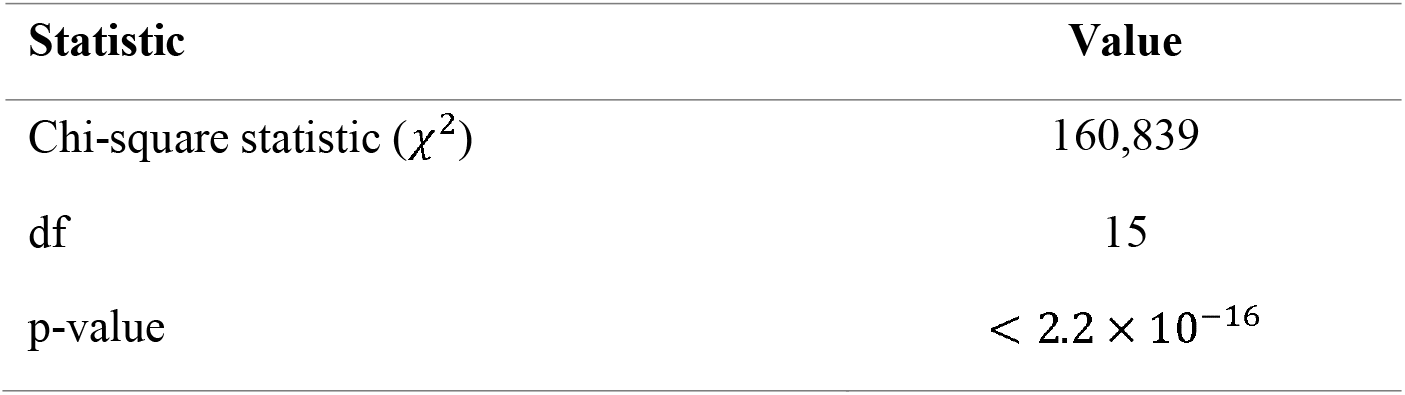
Chi-Square Test for Spatial Heterogeneity.

| Statistic | Value |
| --- | --- |
| Chi-square statistic ($\chi^2$) | 160,839 |
| df | 15 |
| p-value | $< 2.2 \times 10^{-16}$ |

### 3.2 Sensitivity Analysis

Table 3 presents a sensitivity analysis comparing the performance of the primary Bayesian spatio-temporal model with three alternative specifications, using WAIC and DIC. The base model (Spatio-Temporal) yields the lowest values for both criteria, with WAIC at 19121.18 and DIC at 19118.54. The model with a stronger spatial prior yields slightly higher values, with a WAIC of 19121.67 and a DIC of 19120.15. Additionally, using RW2 for the temporal trend further increases the criteria, resulting in a WAIC of 19122.89 and a DIC of 19121.02. Removing the spatio-temporal interaction term also yields higher values: a WAIC of 19122.70 and a DIC of 19120.66. The consistently lowest WAIC and DIC for the base model indicate that this primary specification—with its chosen priors, RW1 temporal structure, and inclusion of a space-time interaction—provides the best balance of fit and predictive accuracy.

**Table 3.** Sensitivity Analysis.

| Model | WAIC | DIC |
| --- | --- | --- |
| Spatio-Temporal | 19121.18 | 19118.54 |
| Stronger Spatial Prior | 19121.67 | 19120.15 |
| RW2 Temporal Trend | 19122.89 | 19121.02 |
| No Spatio-Temporal Interaction | 19122.70 | 19120.66 |

Table 4 presents the posterior summaries for the fixed and random effect components of the Bayesian spatio-temporal model. Parameters are reported as posterior means with 95% credible intervals (CIs) on the log-relative risk scale. Any CI that includes zero is considered not statistically significant. Specifically, rainfall at lags 0 and 3, humidity at lag 0, temperature at lag 0, apparent temperature at lag 0, sea-level pressure at lag 0, and the major rainy season are not statistically significant since their CIs overlap with zero. Rainfall with a lag of one month shows a positive and significant effect (mean = 4.03 × 10^*−6*^, 95% CI: 2.06 × 10^−6^, 6.01 × 10^−6^), as does rainfall with a lag of two months (mean = 2.56 × 10^−6^, 95% CI: 6.64 × 10^−7^, 4.46 × 10^*−6*^). This indicates that a 1 mm increase in rainfall one to two months prior is associated with a small but statistically significant increase in the log-relative risk of malaria. Humidity has a significant delayed effect. A 1% increase in humidity one month prior is associated with a 1.78% increase in risk (mean = 0.0178, 95% CI: 0.0122, 0.0234). The dew point exhibits a more complex lagged relationship, showing a significant negative effect at a 1-month lag (mean = −0.0408, 95% CI: −0.0578, −0.0238), which indicates a 4.0% reduction in risk for every 1°C increase. However, dew point also shows significant positive effects at lag 2 (mean = 0.0082, 95% CI: 0.00176, 0.0146) and lag 3 (mean = 0.0147, 95% CI: 0.00907, 0.0204), corresponding to increases in risk of 0.82% and 1.48%, respectively. Solar energy has a significant positive immediate effect (mean = 0.0634, 95% CI: 0.0265, 0.1003), where a 1 MJ/m^2^ increase is associated with a 6.55% higher risk of malaria. In contrast, the UV Index indicates a significant protective immediate effect (mean = −0.1523, 95% CI: −0.2554, −0.0492), leading to a 15.23% reduction in risk per unit increase. Regarding seasonal indicators, the minor rainy season is associated with a significant 8.06% reduction in malaria risk relative to the dry-season baseline (mean = −0.08399, 95% CI: −0.1410, −0.02695).

**Table 4.** Posterior Summaries for Fixed and Random Effect Components of the Bayesian Spatio-Temporal Model.

| Component | Posterior (Mean (95% CI)) |
| --- | --- |
| ***Fixed Effects*** |  |
| Intercept ($\beta_0$) | 0.072 (0.0153, 0.298) |
| Rainfall $\ell_0$ | $1.59 \times 10^{-6}$ ($-4.78 \times 10^{-7}$, $3.66 \times 10^{-6}$) |
| Rainfall $\ell_1$ | $4.03 \times 10^{-6}$ ($2.06 \times 10^{-6}$, $6.01 \times 10^{-6}$) |
| Rainfall $\ell_2$ | $2.56 \times 10^{-6}$ ($6.64 \times 10^{-7}$, $4.46 \times 10^{-6}$) |
| Rainfall $\ell_3$ | $-2.85 \times 10^{-7}$ ($-2.17 \times 10^{-6}$, $1.60 \times 10^{-6}$) |
| Temperature $\ell_0$ | 0.0066 (-0.057, 0.070) |
| Humidity $\ell_0$ | $7.54 \times 10^{-5}$ (-0.012, 0.0126) |
| Humidity $\ell_1$ | 0.0178 (0.0122, 0.0234) |
| Dew point $\ell_0$ | 0.0191 (-0.016, 0.0543) |
| Dew point $\ell_1$ | -0.0408 (-0.0578, -0.0238) |
| Dew point $\ell_2$ | 0.0082 (0.00176, 0.0146) |
| Dew point $\ell_3$ | 0.0147 (0.00907, 0.0204) |
| Solar energy $\ell_0$ | 0.0634 (0.0265, 0.1003) |
| UV index $\ell_0$ | -0.1523 (-0.2554, -0.0492) |
| Apparent Temperature $\ell_0$ | -0.0236 (-0.0517, 0.0046) |
| Sea-level pressure $\ell_0$ | 0.0167 (-0.00691, 0.0403) |
| Season Major Rainy | -0.00649 (-0.056, 0.0433) |
| Season Minor Rainy | -0.08399 (-0.1410, -0.02695) |
| ***Random Effects Precision*** |  |
| Structured spatial ($\tau_\mu$) | $1.93 \times 10^6$ (0.84, $2.11 \times 10^6$) |
| Unstructured spatial ($\tau_\nu$) | 3.89 (0.72, 9.60) |
| Temporal ($\tau_\gamma$) | $6.22 \times 10^5$ (436.46, $1.20 \times 10^6$) |
| Spatio-temporal interaction ($\tau_\phi$) | 5665.12 (50.42, 37375.97) |

The precision parameters for the random effects confirm a strong latent spatio-temporal structure. The structured spatial effect exhibits high precision (*τ_μ_* mean = 1.93 × 10^6^, 95% CI: 0.84, 2.11 × 10^6^), indicating pronounced spatial smoothing and dependency between neighboring regions. The unstructured spatial effect shows moderate precision (*τ_ν_* mean = 3.89, 95% CI: 0.72, 9.60), capturing region-specific heterogeneity. The temporal random effect demonstrates very high precision (*τ_γ_* mean = 6.22 × 10^5^, 95% CI: 436.46, 1.20 × 10^6^), reflecting a smooth yet well-defined evolution of risk over time. Finally, the significant precision for the spatio-temporal interaction (*τ_ϕ_* mean = 7565.12, 95% CI: 50.42, 37375.97) validates the presence of dynamic, localized risk patterns that vary jointly across space and time.

### 3.4 Predictive Relative Risk of Malaria across Ghana

Figure 1 presents the annual mean estimates of the relative risk (RR) of malaria across the 16 administrative regions of Ghana for the period from 2020 to 2024, as generated by the Bayesian spatio-temporal model. The risk levels are visualized using a graded color scale, categorized into three distinct classes: R1 (low RR: 0–1.0), R2 (moderate RR: 1.0–1.5), and R3 (high RR: 1.5–2.0). The maps indicate a consistent spatial pattern of malaria risk during these five years, with notable shifts in 2024 that highlight changing risk dynamics. The Northern and Northwestern regions, including Upper West, Upper East, Bono, Ahafo, and Western North, consistently exhibited a high risk of malaria, remaining in the highest classification (R3) throughout the entire timeframe. While the Bono East, Oti, Eastern, Western, and Central regions maintained a stable moderate-risk classification (R2) over the five years, the Greater Accra, Ashanti, Volta, Northern, Savannah, and North East regions generally remained in the low-risk category (R1) for most years. A notable temporal occurrence occurred in 2024, with the North East region moving from the low-risk (R1) to the moderate-risk (R2) category, indicating an increase in transmission intensity. At the same time, the Western North region shifted from the high-risk (R3) classification to the moderate-risk (R2) category, suggesting a potential reduction in its malaria burden that year.

**Figure 1.**
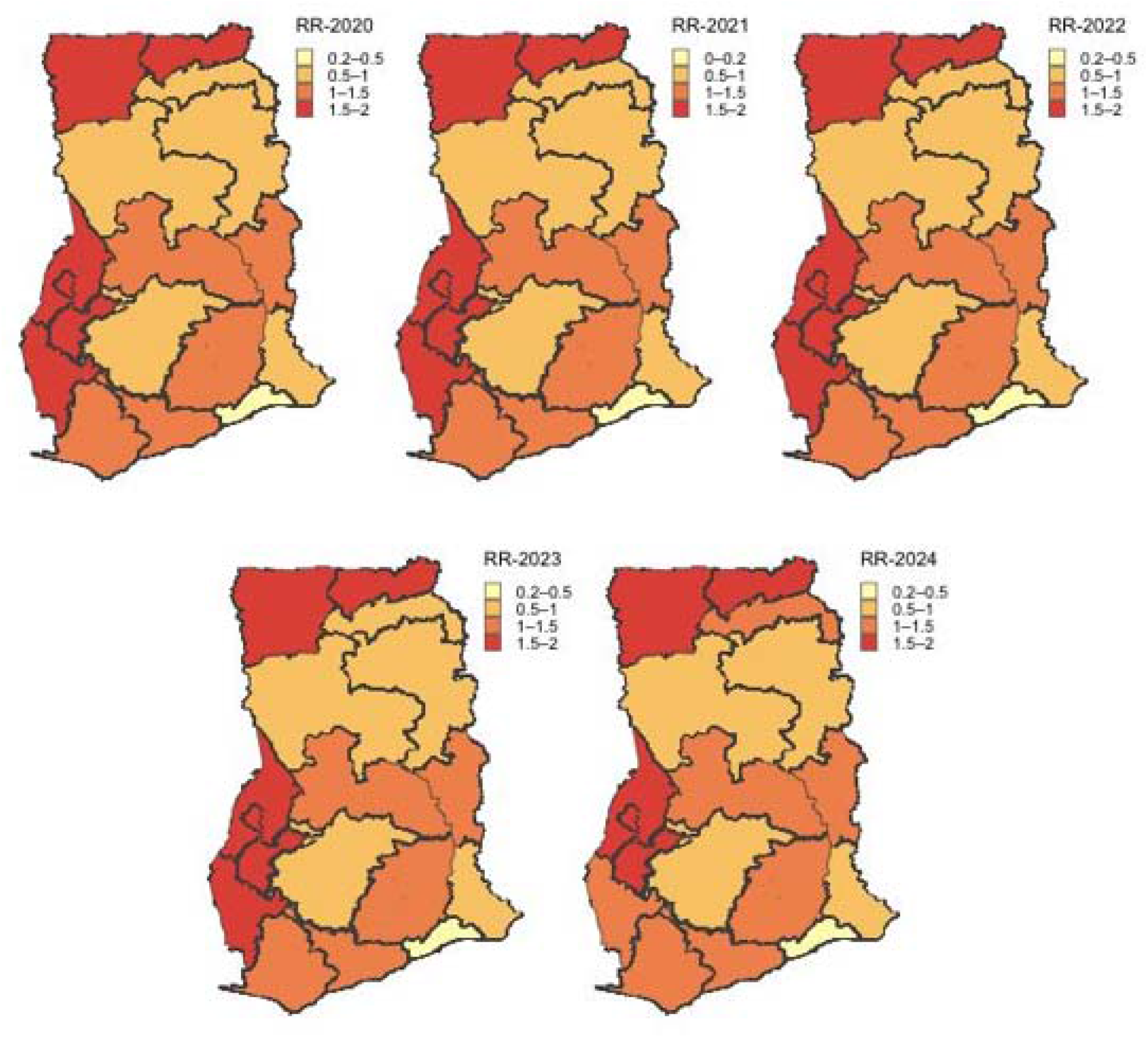
Predictive Relative Risk Maps of Malaria across the 16 Regions of Ghana from the Bayesian Spatio-Temporal Model (2020–2024). The maps show a clear pattern of malaria risk over five years, with significant changes in 2024. The Northern and Northwestern regions, including Upper West, Upper East, Bono, Ahafo, and Western North, consistently faced high malaria risk. Bono East, Oti, Eastern, Western, and Central regions maintained a stable moderate risk, whereas Greater Accra, Ashanti, Volta, Northern, Savannah, and North East regions remained mostly in the low-risk category. In 2024, the North East region shifted from low risk to moderate risk, while Western North moved from high risk to moderate risk.

### 3.5 Hotspot Mapping

Figure 2 presents the annual cluster maps of Local Indicators of Spatial Association, which illustrate the relative risk of malaria across Ghana’s sixteen administrative regions from 2020 to 2024. These maps are generated from posterior estimates obtained via the Bayesian spatio-temporal modeling framework and categorize regions into statistically significant spatial clusters. The classification uses color coding to differentiate between cluster types: high-high clusters are shown in red, high-low in light blue, low-high in navy, and low-low in orange. Areas that do not have statistical significance are depicted in a neutral shade. The analysis indicates a stable spatial clustering structure over time. The Bono Ahafo region consistently forms a high-high cluster from 2020 to 2023, indicating it is a core zone with a significantly elevated, contiguous malaria risk. In contrast, the Ashanti region remains a low-high cluster during the same period, indicating a significant low-risk anomaly within a predominantly high-risk area. The Eastern region consistently exhibits a high-low cluster throughout 2020 to 2024, representing a high-risk area surrounded by lower-risk neighbors. In 2024, a notable shift occurs as the southern coastal regions— Greater Accra and Central—transition to form a low-low cluster, indicating a spatially aggregated area of sustained low transmission. The remaining regions, primarily located in transitional ecological zones, show no significant spatial associations.

**Figure 2.**
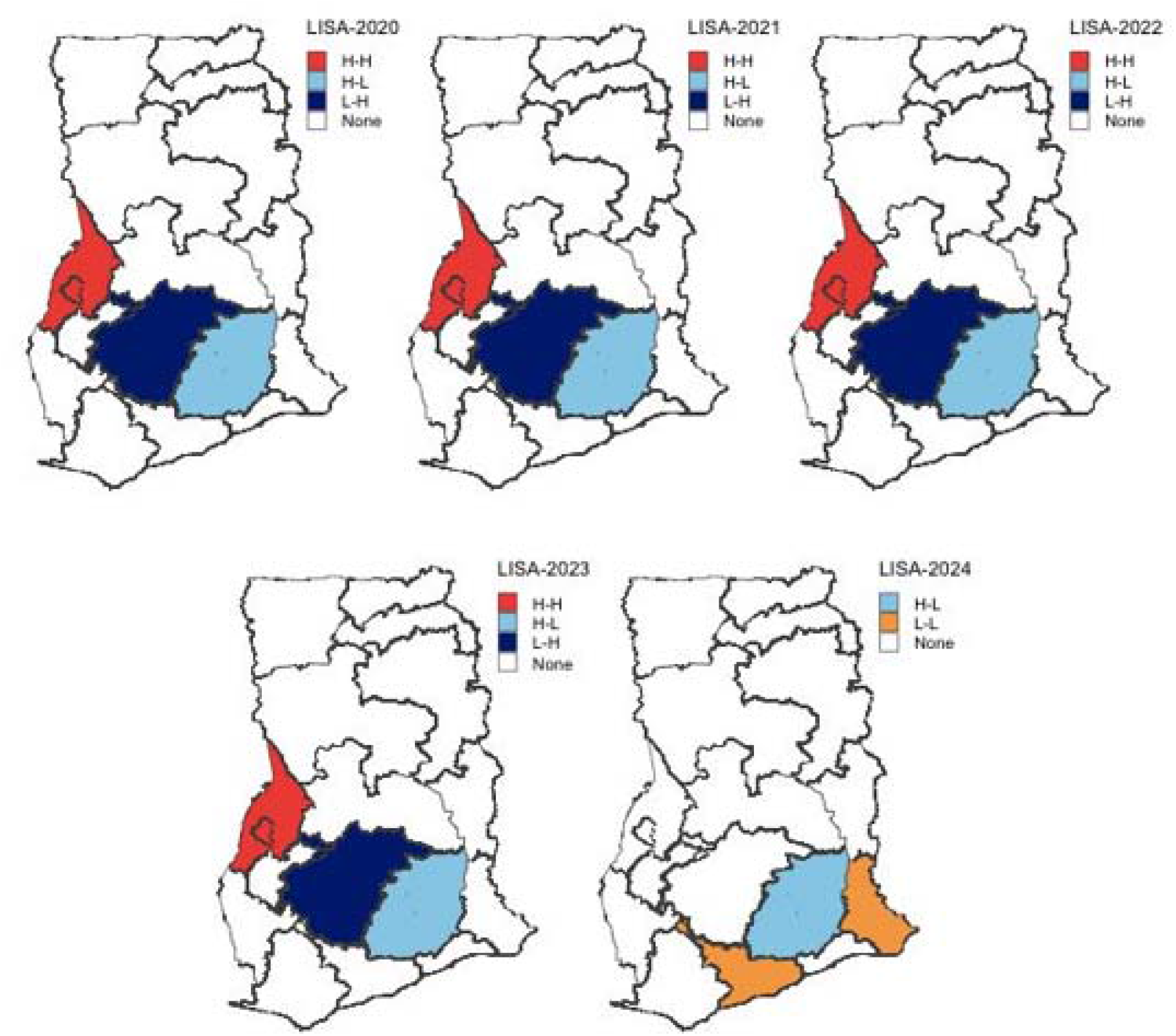
Local Indicators of Spatial Association cluster map depicting significant spatial clustering of malaria relative risk across Ghana’s 16 regions from 2020–2024. The Bono Ahafo region remains a high-high cluster from 2020 to 2023, indicating a core zone of elevated malaria risk. The Ashanti region is a low-high cluster, reflecting a significant low-risk anomaly in a mostly high-risk area. The Eastern region consistently shows a high-low cluster, indicating a high-risk area surrounded by lower-risk neighbors. By 2024, the Greater Accra and Central regions shift to a low-low cluster, indicating sustained low transmission.

## 4 Discussion

This study utilized a Bayesian spatio-temporal model to examine the drivers and patterns of malaria risk across Ghana’s 16 administrative regions from 2020 to 2024. The goal was to identify hotspots with significantly elevated or persistently elevated malaria risk to inform targeted intervention planning and thereby support Ghana’s National Malaria Elimination Program.

Our findings revealed that humidity, measured with a one-month lag, had the strongest effect on malaria risk. Specifically, a 1% increase in relative humidity was associated with a 1.78% increase in malaria risk. This result aligns with existing research indicating that relative humidity above certain thresholds significantly enhances mosquito longevity and biting rates [29, 30]. Atmospheric moisture conditions critically influence vector survival and feeding frequency, which, in turn, affect the intensity of malaria transmission [31]. The delayed effect reflects the continuous need for adequate moisture to sustain adult mosquito populations and ensure frequent human-vector contact [32]. The persistent effects of humidity over one month are consistent with biological understanding, as increased humidity can extend the lifespan of adult Anopheles mosquitoes, increasing the likelihood that they will survive the extrinsic incubation period of the Plasmodium parasite [33].

We also observed a substantial immediate effect of solar energy at lag zero, with a 1 MJ/m^2^ increase associated with a 6.55% rise in malaria risk. While this finding may seem counterintuitive, it is biologically plausible. Increased solar radiation is often associated with higher temperatures and increased evaporation, creating more favorable conditions for larval development by concentrating nutrients in standing water and accelerating the mosquito life cycle [34, 35]. Additionally, the positive effect of solar radiation may be related to rising water temperatures in larval habitats. In contrast, higher temperatures can accelerate development; they may also increase mosquito mortality if temperatures exceed their tolerance limits [36].

Rainfall, measured with a one-month lag, showed a significant positive association with malaria risk, indicating that a 1 mm increase in rainfall one month prior was associated with a small but statistically significant increase in log-relative risk. This finding is consistent with established ecological knowledge of malaria transmission, which indicates that rainfall creates breeding sites for Anopheles mosquitoes. However, the development from larvae to infectious adult mosquitoes requires several weeks [29, 37], which explains the observed lagged effect of rainfall on malaria transmission. The biological rationale for these delayed effects is well-documented; rainfall creates breeding habitats for Anopheles mosquitoes, necessitating time for larval development, adult emergence, and the sporogony of the parasite [38, 39]. This aligns with studies conducted across sub-Saharan Africa that have identified optimal lag periods for precipitation in predicting malaria incidence [17, 20]. Rainfall at a two-month lag also showed a significant positive association, although with a slightly smaller magnitude than the one-month lag effect. The sustained significant effects for two months may reflect the ongoing availability of breeding sites and multiple overlapping generations of mosquitoes following heavy rainfall events [40, 41]. This pattern of peak correlation at one to two months corresponds with previous studies in Ghana that reported peaks in malaria transmission approximately two months after such rainfall events [42, 43].

The study revealed that, with a 3-month lag, the dew point had a significant positive effect on malaria risk, corresponding to a 1.48% increase in risk per 1°C rise. This delayed positive effect suggests the cumulative influence of sustained atmospheric moisture on mosquito survival and the persistence of breeding sites over extended periods [31]. High moisture levels sustained over subsequent months contribute to overall environmental humidity that favors mosquito survival and breeding site persistence [15]. This finding highlights the importance of incorporating longer lag structures in climate-malaria models [7]. Although the two-month lag dew point also showed a significant positive effect, with a 0.82% increase in risk per 1°C, the one-month lag dew point showed a significant negative effect, indicating a 4.0% reduction in risk per 1°C increase. The dew point, which measures absolute atmospheric moisture, is expected to reflect conditions more favorable for mosquito survival than relative humidity alone [31]. However, the observed protective effect at a one-month lag may be due to complex interactions with temperature. For instance, high dew points in combination with elevated temperatures can create conditions that reduce vector survival or alter human-vector contact patterns [34, 44]. The initial protective effect may be linked to overcast conditions and reduced sunlight, which can hinder mosquito flight and activity, leading to a temporary reduction in transmission [10]. This non-linear and time-varying relationship underscores the need for careful interpretation of climate covariates and justifies the inclusion of multi-lag structures in spatio-temporal models. The significant positive effects at both two- and three-month lags, combined with the significant negative effect at one-month lag, reveal a complex, time-varying relationship between dew point and malaria transmission, warranting further mechanistic investigation.

In the current study, the UV Index at lag zero exhibited the most pronounced protective effect among the significant negative associations, corresponding to a 14.14% reduction in malaria risk per unit increase in the index. This finding is biologically plausible, as ultraviolet radiation has direct lethal effects on Plasmodium parasites and reduces the viability of mosquito larvae [30, 45, 46, 47]. Experimental evidence shows that UV exposure disrupts sporogonic development in Anopheles mosquitoes, thereby interrupting parasite transmission [48, 49]. Additionally, UV-B radiation increases larval mortality and prolongs development time, thereby reducing adult emergence [50]. Sublethal exposure also induces oxidative stress and impairs the flight capacity and feeding behavior in adult mosquitoes, all of which lower vectorial capacity [51, 52]. The immediate (lag-0) effect aligns with the rapid onset of UV-induced damage, in contrast to the delayed effects of climatic variables that influence larval habitat availability [30]. These findings emphasize the significance of solar radiation as a determinant of transmission dynamics and suggest that incorporating UV Index data could improve predictive accuracy in spatio-temporal models, particularly in high-insolation settings [46]. Moreover, the protective effect suggests that periods of elevated UV radiation could naturally complement outdoor vector-control interventions [47].

Although the study found that the minor rainy season was associated with a significant 8.06% reduction in malaria risk compared to the dry season baseline, this seemingly counterintuitive finding may arise from complex interactions among rainfall intensity, the stability of mosquito breeding sites, and the timing of interventions, all of which can vary across ecological zones in Ghana [42]. While the major rainy season, characterized by intense and prolonged rainfall, may flush out breeding sites, temporarily reducing mosquito populations [3, 53], the minor rainy season, which typically occurs later in the year, may create fewer sustained breeding habitats or coincide with increased vector control activities following the peak of the major rainy season [54]. This finding underscores the need for models that account for seasonal variability in transmission risk and underscores the importance of timing in malaria intervention strategies [55].

The random effects components, as indicated by the precision parameters, strongly validate the methodological approach of the current study. The extremely high precision of the structured spatial effect suggests that neighboring regions share a significant underlying similarity in malaria risk that is not explained by the included climatic covariates [14, 56]. This “borrowing of strength” is a key advantage of the CAR model, enabling stable estimates even in areas with sparse or volatile data [5, 6]. The Monte Carlo Moran’s I test confirmed the presence of spatial autocorrelation, providing empirical support for incorporating spatial effects into the analytical framework [7, 57]. The moderate precision of the unstructured spatial effect captures unique region-specific factors, such as local healthcare access, bed net usage, or micro-ecological conditions that the model does not account for [58]. The Chi-square test for spatial heterogeneity revealed substantial spatial variation in malaria burden, underscoring the necessity for models that can account for region-specific effects [59].

The current study has also identified persistent high-risk clusters in the northern and northwestern regions (Upper West, Upper East, Bono, Ahafo, Western North), which is consistent with earlier reports of an elevated malaria burden in the savannah and forest-savannah transition zones [21]. The ongoing high risk in these areas may be due to their predominantly savannah ecology, which features a single intense rainy season that creates extensive breeding habitats for Anopheles mosquitoes [40, 60]. The stability of these hotspots over the five years highlights the importance of regional ecological and socio-economic factors that sustain malaria transmission beyond short-term climatic fluctuations [61].

Furthermore, the study identified significant spatial associations: Bono and Ahafo emerged as a consistent high-high cluster from 2020 to 2023, indicating a core zone of elevated, spatially contiguous malaria risk.

In contrast, Ashanti was consistently observed as a low-high anomaly, indicating a significant low-risk area surrounded by higher-risk neighbors, while Greater Accra and Central transitioned to form a significant low-low cluster in 2024, representing a spatially aggregated zone of sustained low transmission. The Ashanti finding is especially noteworthy, as it suggests that, despite being geographically surrounded by higher-risk areas, this region maintains lower transmission levels, possibly due to more effective health system interventions, increased urbanization, or differences in vector species composition. These spatial patterns reveal distinct differences in the geography of malaria risk compared to the geostatistical study by [62], which identified Upper East, Northern, and Ashanti as high-incidence zones based on raw incidence rates. The present Bayesian spatio-temporal model with LISA clustering presents a contrasting view. Bono, Ahafo, and Western North emerge as persistent high-risk clusters that require prioritized interventions—regions that were completely absent from their study. Conversely, Ashanti is reclassified as a low-high anomaly, and Northern falls within the low-risk category, contradicting its earlier classification as high-incidence. These discrepancies stem from methodological differences, including the inclusion of broader climatic variables, formal testing for overdispersion, and the application of spatially explicit LISA analysis within a Bayesian hierarchical framework. The implications for targeted interventions are significant. Resources should be redirected toward the newly identified hotspots in Bono and Ahafo, while avoiding over-investment in areas where raw incidence rates may be misclassified as high risk. These insights, which could be overlooked in purely global spatial analyses, emphasize the value of local spatial statistics for targeting sub-national control measures and advancing Ghana’s malaria elimination strategy.

## 5 Limitations of the Study

The Bayesian hierarchical framework offered reliable estimates of spatiotemporal malaria risk patterns; however, the study has some limitations that should be acknowledged. First, the analysis was conducted at the regional level, which may mask more detailed variations in transmission. Additionally, other environmental factors, such as land use or socioeconomic conditions, that were not included in this study may also influence malaria risk. Finally, the 2020 population estimates were obtained by linear interpolation from projected values for 2021 and 2022, which may introduce inaccuracies in the expected case counts for that year.

## 6 Conclusion

This study used a Bayesian hierarchical spatio-temporal modeling framework to examine the spatial patterns of malaria risk across Ghana’s 16 administrative regions from 2020 to 2024. The model incorporated spatial dependence through conditional autoregressive priors, included unstructured heterogeneity components, and accounted for spatio-temporal interactions. It also addressed overdispersion by using a negative binomial likelihood. The analysis, conducted with Bayesian hierarchical modeling and INLA estimation, confirmed significant spatial dependence and marked heterogeneity across Ghana’s regions. Predictive relative risk maps for 2020 to 2024 showed that high-risk clusters remained persistent in the northern and northwestern regions, including Upper West, Upper East, Bono, Ahafo, and Western North. Regions such as Bono East, Oti, Eastern, Western, and Central maintained stable moderate risk classifications. In contrast, the Greater Accra, Ashanti, Volta, Northern, Savannah, and North East regions primarily remained in the lowest-risk category. Notable temporal shifts observed in 2024 indicated an emerging intensification of transmission in the North East, along with potential improvements in Western North. The spatio-temporal interaction term validated the presence of dynamic, localized risk patterns that changed jointly over space and time. Hotspot identification through Local Indicators of Spatial Association analysis revealed that Bono-Ahafo consistently emerged as a high-high cluster from 2020 to 2023, indicating a core transmission zone requiring intensive, coordinated interventions. The Ashanti region regularly showed a low-to-high anomaly, suggesting the presence of localized protective factors—possibly due to urbanization or differences in health system performance— despite an unfavorable environmental context. The Eastern region maintained a stable high-low cluster, indicating a localized high-risk area that needs targeted interventions, despite lower-risk neighbors surrounding it. In 2024, the southern coastal regions formed a significant low-low cluster, suggesting aggregated spatial suppression of transmission, potentially linked to ongoing control efforts or ecological factors. These findings provide valuable insights for Ghana’s National Malaria Elimination Program Strategic Plan (2024-2028), facilitating targeted resource allocation and climate-informed timing for interventions.

## Author Contributions

**Pascal Antwi:** conceptualization, data curation, formal analysis, investigation, methodology, visualization, writing – original draft, writing – review and editing. **George Muhua:** conceptualization, supervision, validation, writing – review and editing. **Eric Nyarko:** conceptualization, methodology, visualization, validation, writing – original draft, writing – review and editing.

## Acknowledgments

We sincerely appreciate the Ghana Health Service for providing the malaria data used in this study.

## Ethics Statement

As part of the Integrated Disease Surveillance and Response, malaria cases are regularly reported in DHIMS-II. Therefore, no formal ethical approval was required for this study, as we used aggregated, de-identified secondary data for the analysis.

## Data Availability Statement

Relevant data and materials based on which conclusions were made are included in the manuscript. However, the study data are available from the corresponding author upon reasonable request.

